# COST-EFFECTIVENESS ANALYSIS FOR HPV MITIGATION STRATEGIES IMPLEMENTED IN 2018 IN THE REPUBLIC OF MOLDOVA BASED ON INFECTIOUS DISEASE MODELLING

**DOI:** 10.1101/19009886

**Authors:** Andrzej Jarynowski

**Author notes:** © Andrzej JARYNOWSKI.

## Abstract

Human papillomavirus (HPV), is a sexually transmittable virus infection, which is necessary risk factor for developing cervical cancer, first killer in working age women in Moldova. Since 2018 Moldova has modified screening program and vaccination program (mainly externally funded). To assess the performance of the mitigation policy we propose cost-effectiveness analysis according to 2 already implemented strategies. (1) Vaccination of a single age-cohort, although vaccinating a single cohort may not have a substantial effect in other countries with distinct socio-economic situation. (2) Transition to more technologically advance screening ecosystem (changing from Romanowski to Pap smear), which might not necessary be cost-efficient in low resource settings (if GDP per capita will not growth substantially at the same time).

1. We verified that single cohort vaccination is both cost-beneficial (total costs reduction will balance intervention costs around the year 2040) and cost-efficient (with incremental impact in 20 years perspective on the level of 2300 EUR/QALY). Moreover, we found out that single year cohort is more beneficial than 5-years cohort vaccination scenarios in our mathematical model. This behaviour could be explained by a transitional situation in Moldova (HPV epidemic is near outbreak threshold), still small changes of model parameters and initial conditions could cause strong effect in the epidemiology. However, a definitive answer cannot be given with the chosen methodology.
2. Transition between Romanowski -> Pap smear cytology in screening benefits unquestionably in epidemiology e.g. due to higher specificity. However, further maintenance and higher procedure costs could exceed treatment costs, hence intervention costs would gather unacceptable share in whole national limited resources dedicated to public health.

## Introduction

The Republic of Moldova has recently started battle with health problems as sexually transmittable diseases and cancers, but due to financial crisis, lack of program coordination, partly because of a lack of experience, has still high morbidity and mortality. From ECDC (European Centre for Disease Prevention and Control) reports [47] Moldova is a European country with almost highest cervical cancer risks and classified by WHO in the field of sexually transmitted diseases at the level of countries of the third world [56]. Additionally, we observed political transformation and beginning of social norm change in Moldova as increase of sexuality patterns in number of sexual partners (second demographic transition or post-modernist revolution). We would like to model multi-faceted transmission of diseases [2] in the context of determining the best epidemiological control according to budget possibilities. The optimal preventive guidelines are known: cervical screening practice, teenagers vaccination and sexual education. In developed countries mitigation strategies are so successful to think even about disease eradication soon [5]. However, cost-effectiveness of described intervention in resource limited setup is still unknown with high uncertainty in HPV vaccination, screening, and cancer treatment costs.

This paper describes epidemiological and financing trends on HPV and cervical cancer in Moldova according to new program launched in 2018. Around 800k EUR in 2017-2018 was spent on capacity building in new screening technology in cytology and another 600k EUR was spent on vaccination in 2018/2019. We want decision makers to be informed about proposition of changes to be made in the allocation of the health resources required to implement optimal (cost/effective) prevention program. Thus we examine: (1) a single year cohort vaccination scenario to test its conditional cost-effectiveness (possible sensitivity to sexual partner acquisition rates and country economic growth) as well as; (2) cytology change from Romanowski to Pap (Papanicolau) smears, would not necessary be cost-efficient in low resource settings.

### Epidemiology and fact sheets

Human papillomavirus, or HPV, is a sexually transmittable virus infection, which is not only the main, but also necessary risk factor for developing cervical cancer [55]. The time between getting infected by HPV and developing a cancer can be twenty years or more, therefore a dynamic model of human behaviour would be very useful, so that simulations can be made and different scenarios compared. We observe both behavioural change (sexual partner number increases) and demographical change (population ageing) [53]. Among the oncogenic HPVs, the most severe one is type 16, present in about half of all cervical cancer cases. We model one HPV strain (16) and imitate multi-strains environment. Recent studies have shown that the main safety precaution with respect to cervical cancer is going to be a combination of vaccination and screening - since only type specific vaccines (as for type 16 and 18 in our study) are available and there are as many as 15 oncogenic high risk HPVs not covered by vaccine used in Moldova.

The Republic of Moldova is one of Europe’s poorest nations (whenever we use statistics of Moldova, we mean right Nistru bank only). Total yearly expenditure on health amounts is limited to just 150EUR per capita and reliable data on cervical cancer is missing. Some official statements are untrustable due to well known corruption mechanism of reporting “virtual” patients [28]. The economic situation over the past 2 decades has not allowed for health systems development [57]. Till now some statistics differ significantly dependently of data source. Moreover, public health system in Moldova had no capacity to couple with official screening program till 2017 (women >20 y. o. every 2 years), where more than 700k test should be done yearly (in 2014 there were around 200k unique cytologies and many sites were already overloaded). Demographically, Republic of Moldova had (in years 1998-2014) a population of 1.4-1.6 millions women ages 15 years and older who are at risk of developing cervical cancer. Sexual active woman cohort (age 15-64) was in range 1.25-1.35 millions [45]. We observe both behavioural change (sexual partner increase) and demographical change (population ageing, but still very young), which both corresponding to second demographic transition since Soviet Union collapse. Last 10 years estimates indicate that every year 400-550 (approx data) women were diagnosed with cervical cancer and 145-220 (register data) died from the disease [22]. Estimated societal cost was around 5000 disability-adjusted life years (DALYs) every year (or 3000 QALY quality-adjusted life years) [10, 50]. Cervical cancer ranks as the 2nd most frequent cancer among women in Republic of Moldova and the 1st most frequent cancer among women between 15 and 44 years of age with share of 39% of all kind of working age woman cancer cases [9]. The incidence of cervical cancer, had increased from 2005 to 2009. Data on the HPV burden is not available, but generalizing old studies with data from other south-eastern European countries, 15-20 % of women in the general population have HPV and 80-90% had it in their live [54]. Prevalence of the most oncogenic HPV-16/18 strains is estimated on 2-5%.

Moldova introduced HPV vaccines in selected subpopulation in 2016 and full coverage of 10 y. o. girls with support from Gavi in 2018 only [Fig. 4]. First national wide suboptimal screening program was introduced in 2013 and updated in 2016 to optimal age-range in almost similar schema as we proposed a year before [61]. The majority of cervical cytology was processed by Romanowski staining technique – standard for the former Soviet Union [52].

**Fig. 1.**
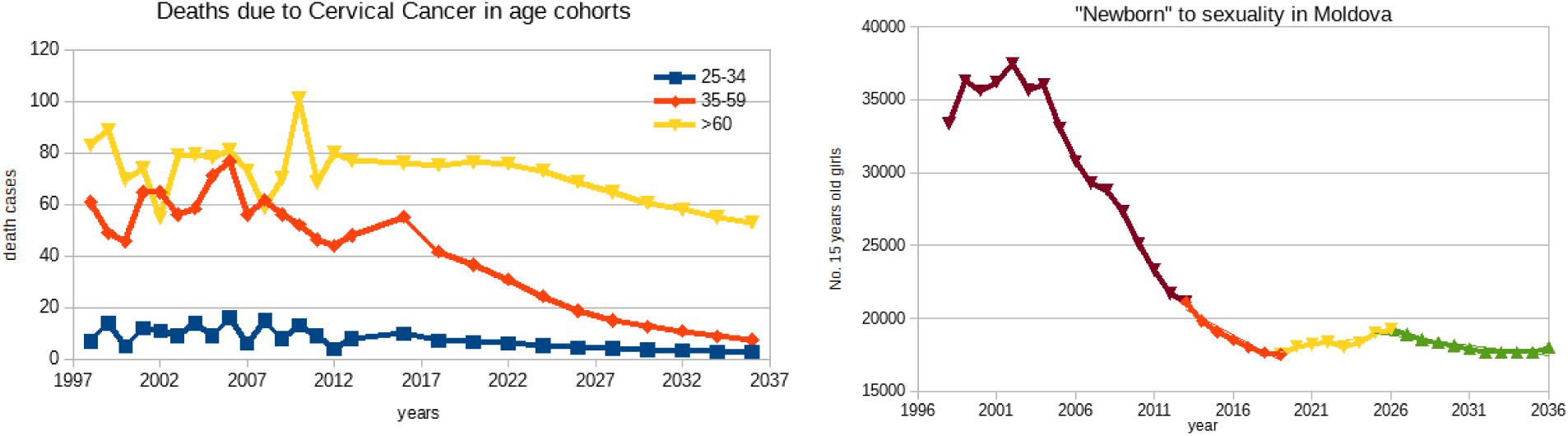
Selected demographical historical data till 2016 and projection (averaged) till 2030. Cervical cancer death cases in age cohorts. {left} Number of 15 years old girls entering sexual maturity age {right}. Source: Author’s calculations based on the Institute of Oncology and Statistics Moldova data.

**Fig. 2.**
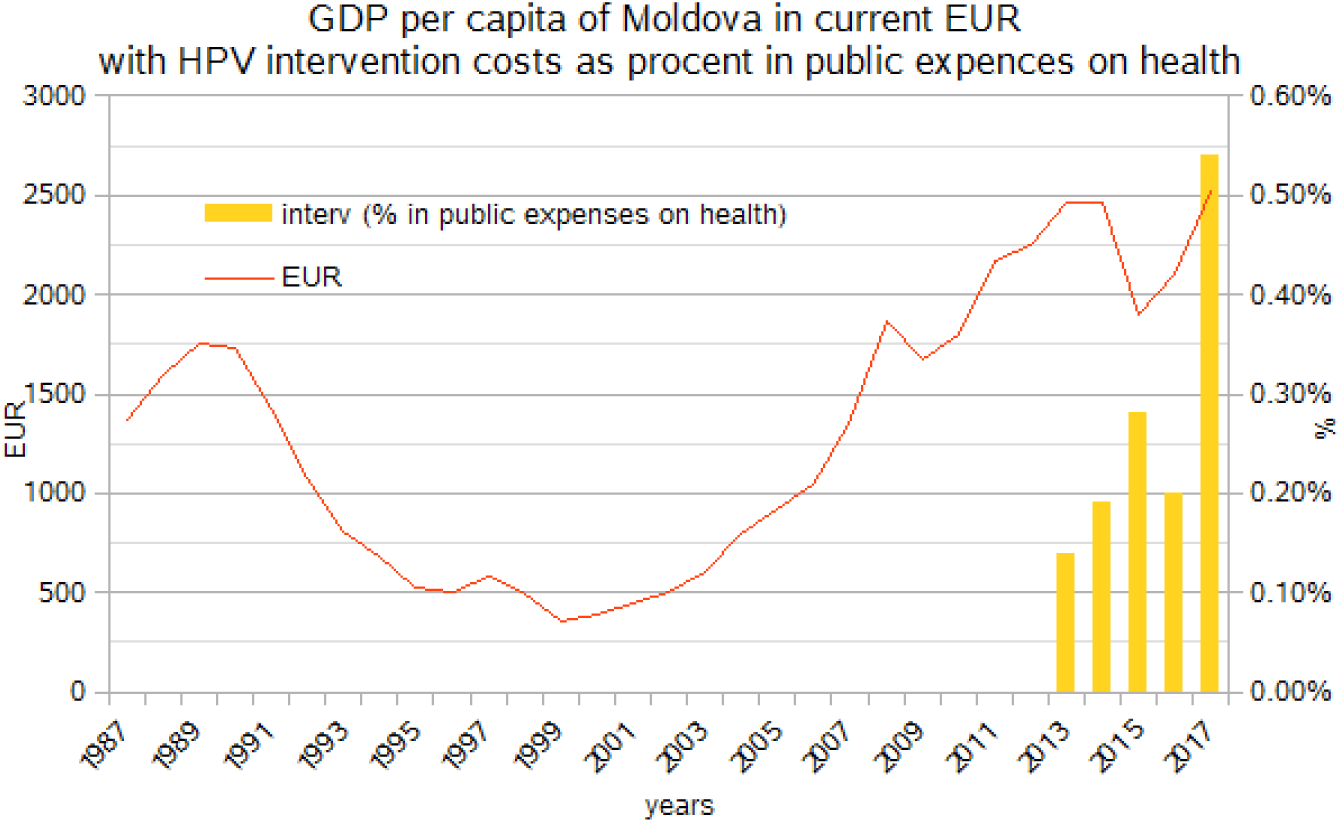
Moldovan GDP patterns (with unpredictable political and economic situation) and HPV intervention costs – the third health expenditure -as % in total health expenses (in comparison Australian intervention in HPV costs are 8 times higher than Moldovan and the same time consume 5 times less available resources) Source: Author’s visualization based on the Statistics Moldova data.

**Fig. 3.**
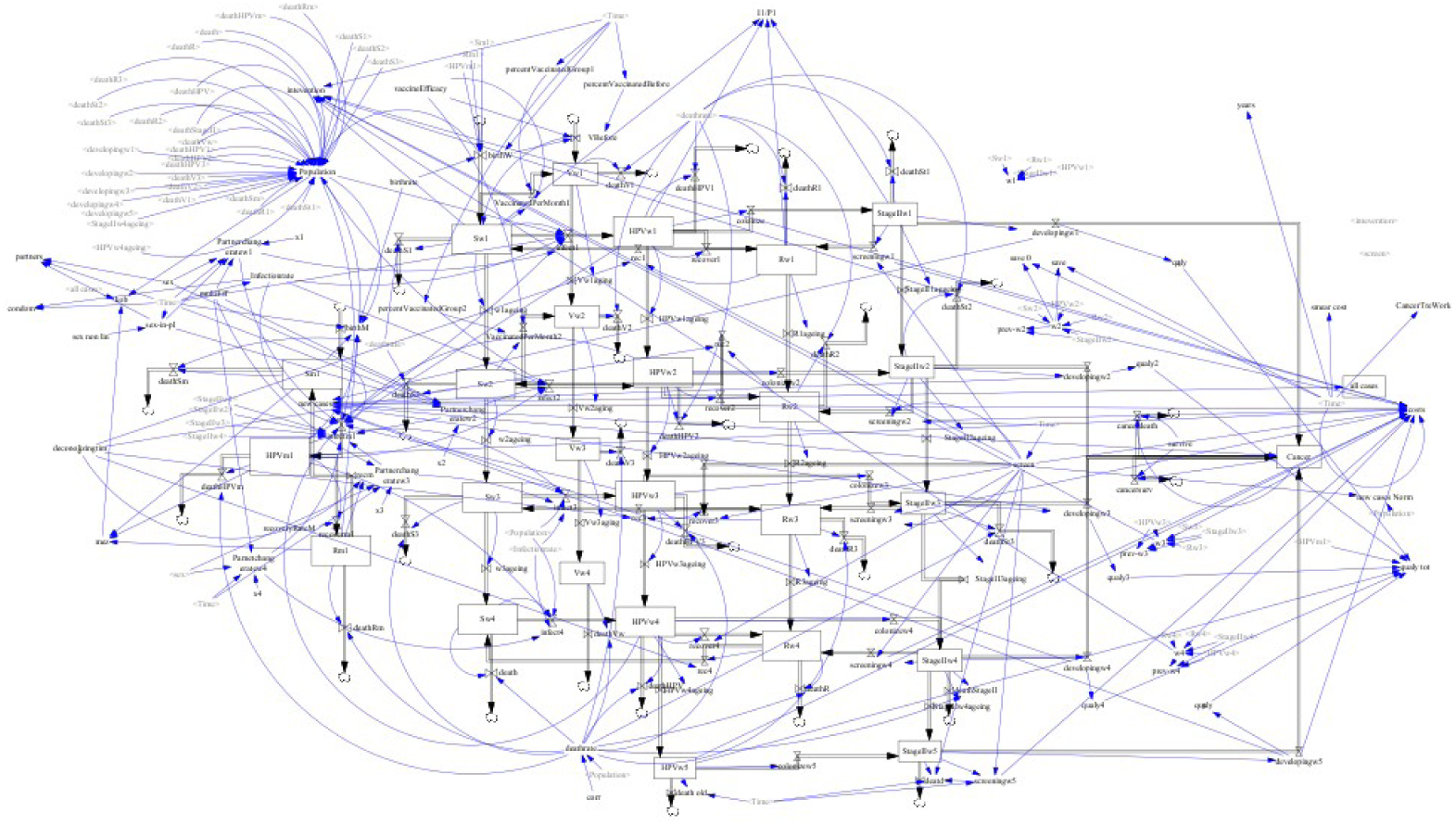
Visualization of this very complicated model in Vensim (Exportable version can be obtained from Github: https://github.com/ajarynowski/HPV_Moldova – free Vensim reader is available from www.vensim.com) Source: Author’s own model available on Github

**Fig. 4.**
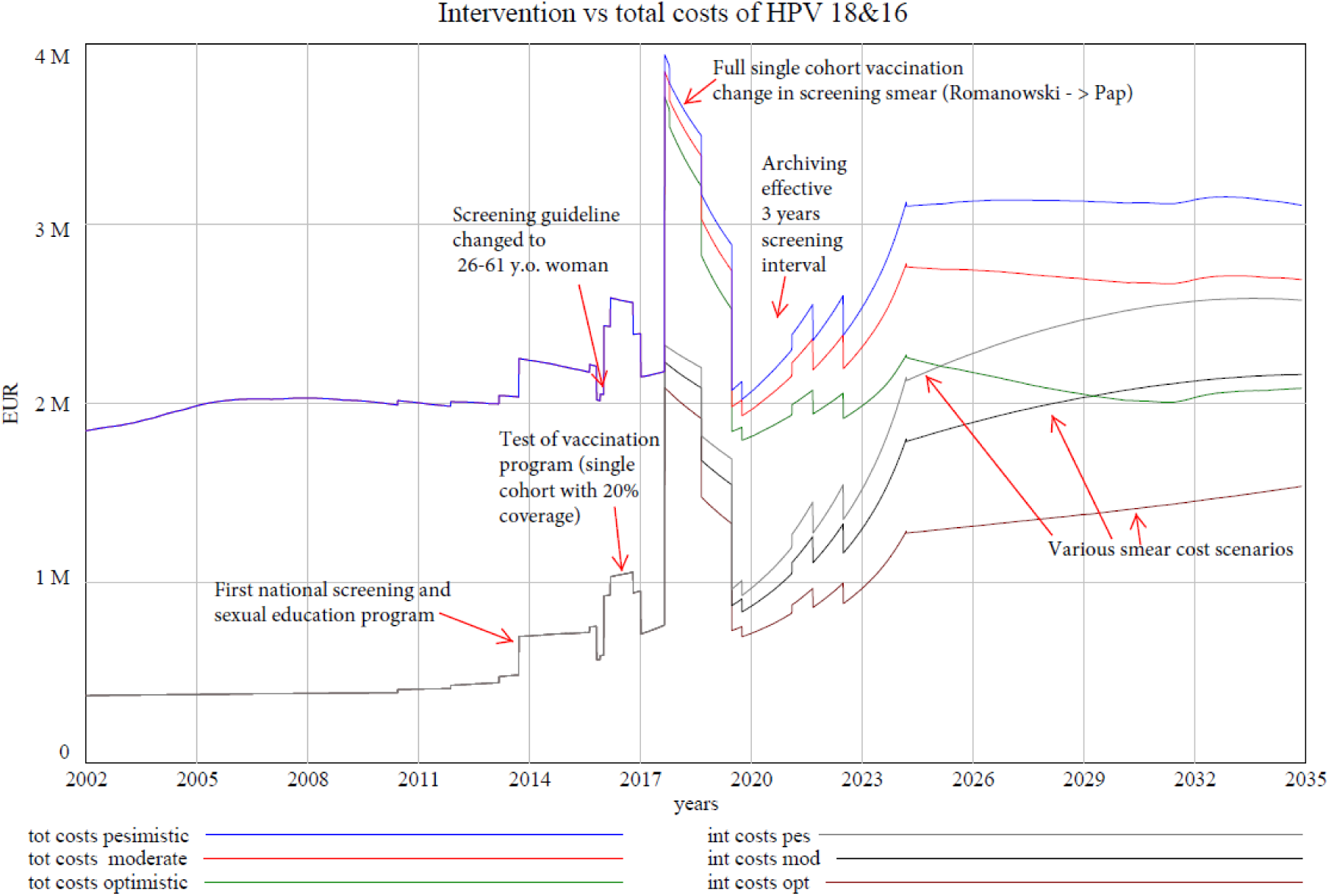
Simplified Moldovan history of pathogenic HPV strains costs. Intervention (int) costs (sexual education, vaccination, screening) and Total (tot) costs (intervention costs with mainly cervical cancer treatment as well as other treatment costs of pre-cancer abnormalities, anus cancer, genital warts) Source: Authors own projections.

### Cost-effectiveness analysis

Let consider more precise monetary and populational effects of intervention used in health economy [8]. To do so, we introduce quality-adjusted life-year (QALY) - a measure of disease burden, including both the quality and the quantity of life lived. The QALY is based on the number of years of life and adjusted to health state (where 1 is a perfect health and 0 is death) that would be saved by the intervention. QALY indicates in the best way the benefits of intervention, because it is sensitive to current patient age (literally to current life expectancy). The intervention is highly cost– effective if its incremental cost per QALY yearly [38] is below GDP per capita of given country (∼2.5k EUR for Moldova) or it could be considered as cost–effective if its below 3*GDP (∼7.5k EUR for Moldova) [58]. In previous papers we showed, that targeted vaccination is cost-effective (the incremental QALY between targeted and no vaccination is less than 4k EUR per QALY in 40 years perspective, however practical implementation could be very difficult), while screening too old and too young woman is not cost-effective (the incremental cost per QALY between national and our scenario was 20k EUR per QALY in 20 years perspective) respectively to Moldova GDP [61]. It is important to mention, that cervical cancer incidence probably will be decreasing any way in Moldova - even with no new additional interventions [29]. Moreover, Moldovan cervical cancer perspective [Fig. 1, 7] looks much better than in other Eastern and Central European countries [1], probably due to recent transition of screening guidelines from opportunistic to organized in beginning from XXI century [12], because of relatively young society and increasing knowledge of STI (sexual transmittable infections).

**Fig. 5.**
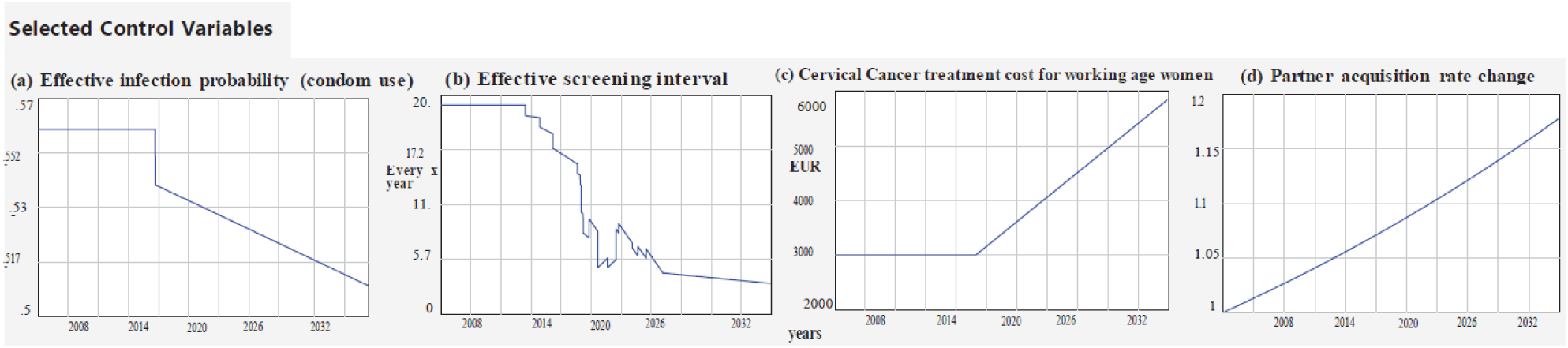
Selected control variables in model projection ((a) effective infection probability – condom use, (b) effective screening interval in years, (c) cancer treatment cost in EUR, (d) sexual partner change rate as multiplicator) Source: Author’s own assumptions.

**Fig. 6.**
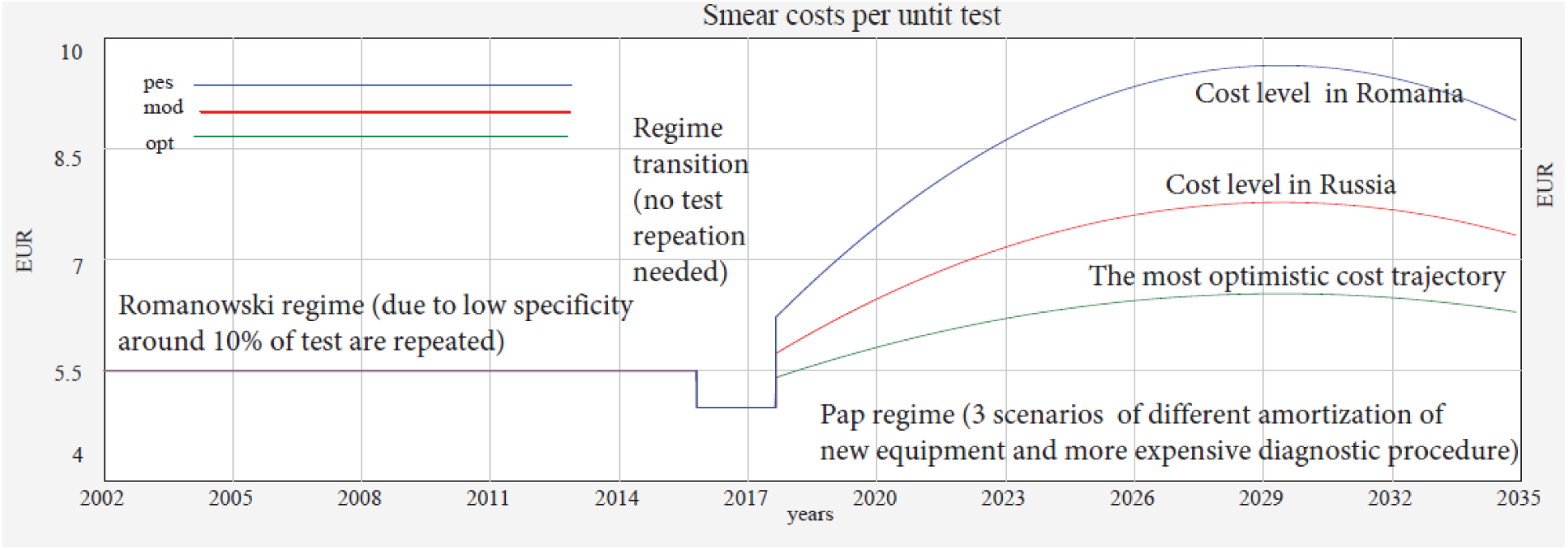
Smear cost per unit for pessimistic (pes), moderate (mode) and optimistic (opt) scenarios. In transitional time window (2017-2018) cost per procedure (operational cost) deceased, because capacity building costs ∼800k EUR (training old and hiring new staff, laboratory equipment etc.) includes partially operational costs. Source: Author’s own assumptions.

**Fig. 7.**
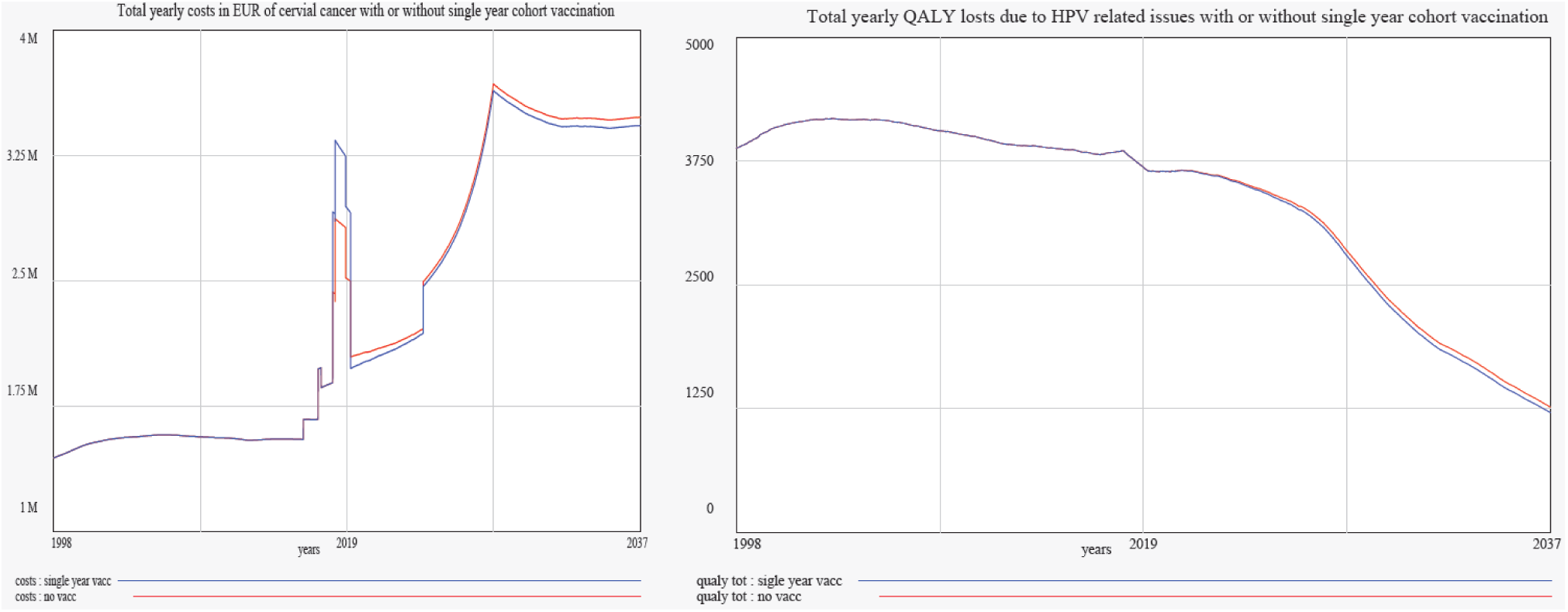
Comparison of total costs of HPV related issued (interventions, genital warts and cancers) {left} and QALY losses {right} with and without vaccination in 2018. Source: Author’s own projections.

### Research Questions

Moldova has modified since 2016 screening program [52] almost the same as we recommended [61] with small difference in maximum age of screening with 61 instead of 65 which could be an artefact of our age-cohorting schema. Other analytical research approach as PRIME [32] and OPTIMA [59] have provided sufficient cost – effectiveness analysis of standard procedures as screening and vaccination, however their universal tools (both of them were built to be used in many counties) do not fit Moldovan specificity in many details. Big intervention in the period of 2017-2020 with main objective as reorganization of screening and new vaccination was externally funded in amount of 1.2M EUR [48]. Ministry of Health declared the will to continue vaccination program already for years 2019-2025 [60], but we are not sure about real financing this project (money in amount of 400k EUR are secured for 2019 only [24]). There is also a strong disbelieve in Moldovan society about vaccinations safety issues [49] with strong anti-vaccination political fuel [44]. There is still an open question about cost effectiveness of point (single cohort) vaccinations, because vaccinating single cohort could not have no satisfactory effects for whole population [39]. The real scenarios assuming vaccinating 10 y.o. girls in 2018 was not implemented by us, because the first effects would be visible too late for simulation time span. We verify instead a single year vaccination of 14 y.o. girls in 2018, so we can keep projection of costs, demographics and epidemiology for next 20 years only. Quadrivalent vaccine (targeting HPV oncogenic strains 16, 18, and also non-oncogenic stains 9 and 11 which cause genital warts) is used in 2018, so we introduced additionally to previous studies, an effects on genital warts (cost of curing and QALY losses). Increase in cytology cost was also updated due to change from Romanowski to Pap smear. Vaccination in a single action a single cohort has not been considered from health economic perspective for Moldova yet. Moldova case cost-effectiveness analysis is also important to be disseminated worldwide [13], due to similar externally founded projects in other Low and Middle Income countries (e.g. GAVI-the Vaccine Alliance).

Research question 1: Testing possible impact of new ‘single’ 2018 year project funded mostly by external sources - vaccines for cohort of girls born in 2007 ∼600k EUR (400k EUR is a financial cost for program continuation in 2019 with GAVI substitution):

- Estimating incremental costs of adding HPV vaccine in single 2018 year to the existing immunization program (reasonable assumption that Moldova will not continue full coverage vaccinating because of lack of resources);
- comparing scenarios of “single year vaccination” in full coverage in 2018 against vaccination of the same amount of girls in the same age but in 5 year time span (20% of each cohort each year). Research question 2: Testing best budget possibilities according to national screening guidelines since 2018 (Romanowski ->Pap cytology change [41]) with capacity building [52] cost of 800k EUR (also funded mostly by external sources):
- Comparing scenarios of various procedure cost per unit (pessimistic – with Romanian price in long term, moderate with Russian price [14] in long term, and optimistic – no significant change).

### Model and results of adjusting realistic scenarios

According to provided information, model for Moldova was proposed. It is a set of deterministic differential equations (implemented in Vensim). Stochasticity [18] was introduced in sexual partner matching schema. The model has aggregated the most important path of infection (heterosexual contacts only), cancer development and prevention scenarios (more than 100 equations and 200 parameters). HPV dynamics (Transmission dynamics via sexual contacts) associated also with the occurrence of cervical cancer (cancer development) was inspired by Polish model implementation [26]. Mathematical formulation of HPV related issues have been already carefully analysed, because its epidemiology has been widely described and modelled in recent years [6, 42]. We have prepared cost-benefit and cost-effectiveness analysis for various vaccination strategies, various screening programs with control over other preventive programs (using condoms/sexual education) for Moldova, based on its own demography [46] and sexual behaviour of heterosexual part of population only [40].

We used data since 1998 (2002) to 2014 (2017) to adjust model parameters and we project till our result around 2035 (2038). We use mathematical [19] and sociological concepts [12] within complex system methodology. Mathematical modelling of infectious diseases transmitted by sexual contacts (as HPV) is increasingly being used to determine the impact of possible interventions (there are dozens of such studies in literature [25, 26, 43]. We have used probability of infection per a new partner as a main transmission driver. The shape of sexual partner distribution [34] was followed from Finish study [3] and adjusted by scaling to obtain mean partners number in Moldova. The most unknown demographical parameter is the increase in partner numbers [35]. That increase of partner’ acquisition was introduced as a modifiable variable and tested for few scenarios [29]. In our model woman are stratified in 5 years cohorts (stocks) starting form 15 y. o. Effective screening intervals (average interval between smears) were previously implemented as a variable (more important from cost/effectiveness analysis) describing healthcare system capacity [29]. Change of screening procedures from opportunistic to regular is also represented in effective screening frequencies [9] and costs [Fig. 4]. Condom use was also tunable variable, while sexual education significantly increased in recent few years [35, 51]. Standard model is better described in extended version of previous report [27], and new functionalities are mentioned directly in this paper. Model and its parametrization in Vensim environment is publicly available: http://github.com/ajarynowski/HPV_Moldova

In our calculations we provide economic costs (total expenses), which differ from financial costs (Moldova Government expenses), because of importance of foreign aids. Intervention costs (sexual education, vaccination, screening) till 2017 counts as ¼ of treatment costs. In big investments in Moldovan Cervical Cancer Program in years 2017-2020 intervention costs (funded by GAVI, World Bank, United Nations Population Fund-UNFPA, Swiss Contribution -SDC, European Union -UE, etc.) exceeds treatment costs.

### Results - Research Question 1) Romanowski->Pap smears

There is only a little difference in epidemiology (specificity) between Romanowski and Pap smear in specificity [Tab. 1]. Lower specificity of Romanowski can be corrected by repeated Pap test (mainly with borderline results) [52].

**Tab.1.**
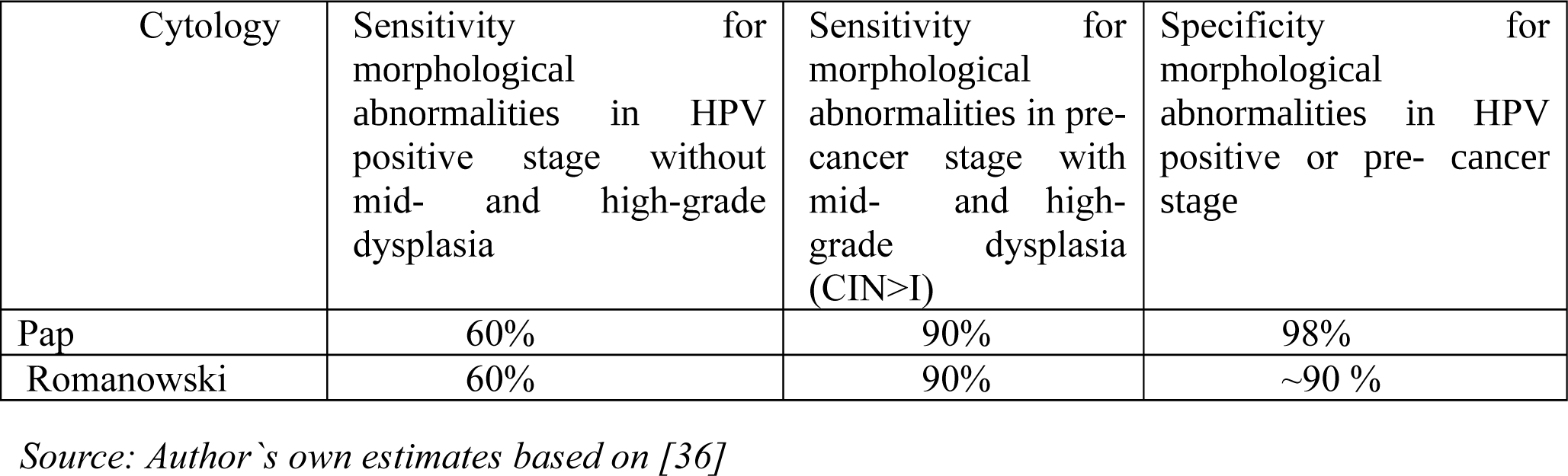
Characteristics of smears. False Positive rate change-> test repetition (up to 10% due to unclear result)

However there is a substantial difference in cost procedure and Pap smear could be 30% to 90% more expensive for a single procedure [Fig. 6], but the same time number of procedures would decreased by around 10%, because there is no need for repetition any more.

While the rising trend in innovative medical technologies (as Pap smear in this case), the chances for efficiency gains and costs reductions are usually large. However, new technology-enabled models of healthcare delivery are not always cost-effective. Capacity building (transitional) costs in this case were covered by foreign sponsors, but equipment amortization and staff cost should be sustain by Moldovan themselves. We try to anticipate costs per procedure in next 20 years [Fig. 6]. We choose target level of future price from Romania (as a pessimistic scenario), Russia (moderated scenario) and current price with most likely interest rate (as optimistic scenario). We assume 15 years amortization period for equipment (characteristic convey curve shape) [Fig. 6]. Reader must be aware, that currently PCR test together with liquid base Pap smear is a standard in developed countries, however machine learning development in image processing for low-cost diagnosis [17] could change whole landscape in next years.

Let’s consider baseline scenario, for which Moldova will stay in Romanowski regime. If we assume that drop-out rate at the level of 20% in repetition tests, there will be around 12 QALY/yearly loses more that in Pap scenario in 20 years projection. Around 10 cancer cases and 4 deaths could be averted with the transition to Pap in 20 years perspective. However, cost comparison is more difficult due to potential instability of Pap procedure price [Fig. 6], so incremental cost-effectiveness vary from 0 in optimistic to 50k incremental EUR/QALY in pessimistic scenario.

### Results - Research Question 2) ‘Single’ cohort vaccination

First of all, we compare single cohort vaccination against no widespread vaccination at all. Vaccination is both cost-beneficial (total cost reduction balance intervention cost around year 2040) and cost-efficient (with incremental impact in 20 years perspective on the level of 2200 EUR/QALY) [Fig. 7].

Thus we examine such a single year vaccination scenario to show its conditional cost-effectiveness in comparison with 5-year cohort vaccination scenario. We assume that 15120 girls has been vaccinated for both single year cohort and 5-year cohort scenarios two years after and before 2018 [Fig. 8].

**Fig. 8.**
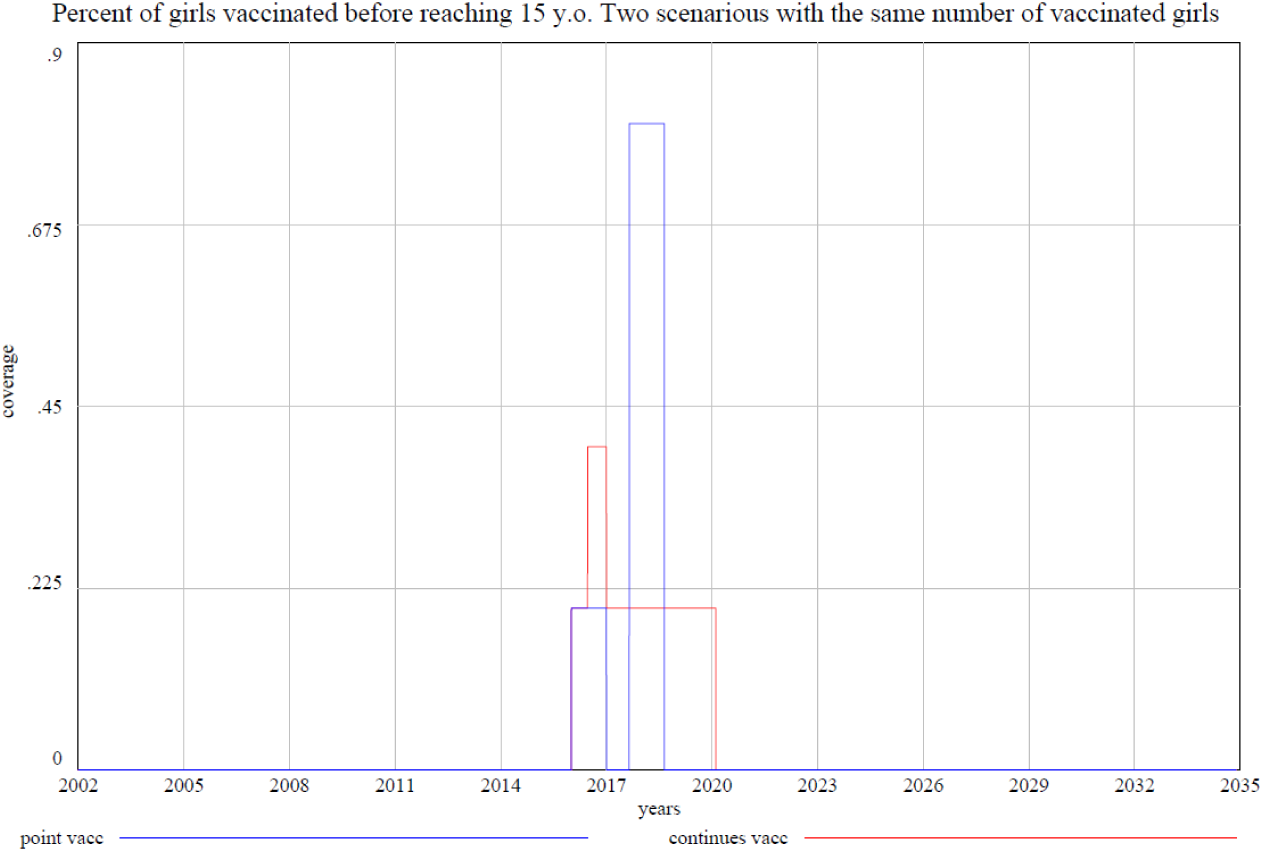
Vaccination coverage of 15120 girls in both scenarios single year/5-years cohort. Source: Author’s own assumptions.

We found out that difference in number of HPV infections is ∼610 and difference in QALY saved is ∼ 80 in favour of single year against 5-years cohort vaccination in next 20 years.

Comparing single year with 5-years cohort in young woman (15-19 y. o.) we observed the increase of infection number between single year and 5-years cohort scenarios in first 1-2 years (because we started 2 years earlier vaccination in 5-years scenario), then around 2019-2022 there is a significant reduction in infections, and there is no long term difference between scenarios (due to aging – moving vaccinated girls to older cohorts). In males, first difference between single year and 5-years cohort scenarios appears in next 5-10 years (when vaccinated women their early 20th years old - the most sexually active period of their life [18]).

### Conclusions, Recommendations and Limitation

The optimal preventive guidelines for cervical cancer are known [42]: cervical screening practice, widespread vaccination and sexual education. However, interventions with the highest impact and lowest price should be prioritized [33]. In addition, Moldova – lower middle income country – straggles with challenging demographic processes, like the ageing of the society [15, 21] and the increase of sexual activity [16] combined with the unstable economic situation [37]. We suggest (according to our simulation) that the official preventive program for cervical cancer in Moldova is optimal in terms of costs and medical efficiency in vaccination schema and suboptimal in screening protocols (optimal in medical efficiency only). We present a simplified representation of the system with around 200 static parameters and additional 20 time dependent control parameters – variables (such as condom use, effective screening interval, partner’s acquisition rate, cancer treatment cost [Fig. 5], etc.,). We consider different scenarios varying the tunable variables (vaccination coverage type [Fig. 8] and smear cost [Fig. 6]). Exact parametrization of the model used for this paper can be found in repository (github). However, reader must be aware, that presented model and parametrization has been chosen according to a heuristic methodology [31]. Although we explored other models and parameters configuration, we cannot claim, that our model is the only and the best one for targeting research question 1 (vaccination) and 2 (screening). On the other hand, the main advantage of our model in comparison with well-established modelling tools already applied to Moldova (developed by WHO – PRIME [32] and sponsored by World Bank OPTIMA [59]) is its flexibility. In our model, we can test different scenarios (as cohort vaccination, and smear price dynamics), which is not possible or feasible in PRIME and OPTIMA, and our findings should be understood as complementary results only.

The idea of transition from Romanowski to Pap smear cytology (research question 1), is unquestionable (due to higher specificity of the last). However, further maintenance and higher procedure costs [Fig. 6] might exceed treatment costs, implying unacceptable share in whole national limited resources dedicated for public health for intervention costs [Fig. 2, 4]. We found an interesting paradox, that transition to more technologically advanced health system (changing from Romanowski to Pap smears), would not necessary be cost-efficient (incremental cost-effectiveness from 0 to 50 k EUR/QALY) in such low resource settings as in Moldova and unstable GDP growth perspective. Reader must be aware, that we simplified difference between Romanowski and Pap smears in procedure cost (trained staff and lab equipment amortization) and specificity (test repetition in Romanowski method) issue only, even both methods have many variants.

We reopen discussion about vaccination guidelines in low-income countries (as Moldova), where cost of widespread action could be too high for local governments (research question 2). Vaccination could be both cost-beneficial (total costs reduction balance intervention costs before 2037) and cost-effective (with incremental impact in 20 years on the level of 2200 EUR/QALY). Moreover the single cohort (point) vaccination (as it was introduced in Moldova) exceeds the 5 years cohort by approximately 610 less infections and approximately 80 less QALY lost in 20 years’ time horizon. The possible explanation of this nonintuitive behaviour might be because HPV in Moldova is rather close to epidemic reproduction threshold rate, still small change of model parameters and initial conditions could cause strong effect in epidemiology. Main effect of intervention is probably via men, which avoid infection (mainly around year 2025 – the peak of sexual activity of vaccinated girls) and will not infect other women. The decrease in infection numbers (and QALY loses in consequence) resulting from the ‘single’ teenage cohort HPV vaccination in 2018 might provide protective effects in heterosexual men through ‘local’ herd immunity [11]. This can have an effect probably while changing partners in Moldova is still not as common as in other countries [7, 23]. However, change in initial conditions and parameter values could diminish positive effect (e.g. higher partner’s acquisition rate). Another limitation is System Dynamic approach, so few features as real sexual patterns matching were not implemented. There is no distinguish of rural and urban population or any spatial resolution [4] in the model, which have impact both on partner’s acquisition and vaccine/screening uptake. Phenomenon of single cohort vaccination cost-effectiveness must be investigated more carefully in different methodology (as ABM Agent Based Modelling) from presented one (System Dynamics) to understand role of men in immunisation efficacy for example. Moreover, no proper sensitivity analysis could be readily performed using the current methodology (limitation of Vensim software) in presented study, even we have found dependence on changing condition, so only through ABM - obtained results could be generalized. Another limitation is time horizon for 20 years, which is already too far for price/demography/technological assumptions and too short to even test efficiency of vaccination of cohort of 10 y. o. girls (so we could test vaccinating 14 y. o. instead). We have used aggregated age cohorting with 5 years window, which on the one hand side is a popular approach [3, 20] – on the second hand it has significant limitations, so year by year analysis could be more proper for single year cohort vaccination schema.

We recommend continuation of vaccination which is both financial (with GAVI substitute) and economic cost-effective. In terms of screening technology transition: there is no way to go back, however screening costs have to be periodically monitored and national guidelines could be revisited (if necessary) according to economic situation in Republic of Moldova.

## Data Availability

Model (Vensim), data (e. g. Statistics Moldova, Institute of Oncology) and projection available at github repository

http://github.com/ajarynowski/HPV_Moldova

## Acknowledgment

AJ would like to thank to Liana Cernov, Florentin Paladi, Ghennadii Gubceac from Moldova State University, Vitaly Belik from Free University of Berlin, David Regan, Trevor Dougherty and Richard Gray from Kirby Institute in Sydney as well as PANTHER project for partial financial support. Codes are publicly available in a repository: https://github.com/ajarynowski/HPV_Moldova

## Notes

### Competing Interest Statement

The authors have declared no competing interest.

### Funding Statement

partly funded by PANTHER (EU scholarship)

### Author Declarations

All relevant ethical guidelines have been followed and any necessary IRB and/or ethics committee approvals have been obtained.

Any clinical trials involved have been registered with an ICMJE-approved registry such as ClinicalTrials.gov and the trial ID is included in the manuscript.

